# Impact of depression on treatment progression in type 2 diabetes: A UK retrospective cohort study using the Clinical Practice Research Datalink Aurum database

**DOI:** 10.1101/2025.11.19.25340563

**Authors:** Alexandra C Gillett, Dale Handley, Renu Bala, Katherine G Young, Jess Tyrrell, Cathryn M Lewis

**Author notes:** Corresponding author: Alexandra C Gillett, Social, Genetic and Developmental Psychiatry Centre, Institute of Psychiatry, Psychology and Neuroscience, King’s College London, London, UK.

## Abstract

**Aims:** To investigate the association between depression history, timing, and progression of diabetes treatment in type 2 diabetes (T2D).

**Methods:** We conducted a cohort study using primary care records from the Clinical Practice Research Datalink Aurum database (2011–2023). Adults with T2D initiating oral glucose-lowering monotherapy (index) were included. Depression history, identified using clinical codes, was categorised by the most recent code before index: recent (≤1.7 years), intermediate (1.7–12.8 years), distant (>12.8 years), or none. Outcomes were time to treatment intensification (adding or switching drug class) and insulin initiation. We used Royston-Parmar survival models to estimate odds ratios (ORs) and 95% confidence intervals (CIs), adjusting for demographic and clinical variables.

**Results:** Among 378,935 included individuals, 25.1% had a history of depression. Compared with no prior depression, recent depression was associated with higher odds of treatment intensification (OR 1.20, 95% CI: 1.17-1.23) and insulin initiation (OR 1.29, 95% CI: 1.23-1.36). Intermediate and distant depression were also associated with higher progression-odds, though more modestly.

**Conclusions:** Depression, particularly recent episodes, is associated with earlier treatment progression in T2D, highlighting the importance of integrating mental and physical healthcare in diabetes management.

## 1. Introduction

Type 2 diabetes (T2D) is a common, growing and complex cardio-renal-metabolic disorder [1]. In the UK, the prevalence of T2D increased from 4% in 2011/12 to 5.8% in 2021/22, contributing to a significant diabetes healthcare burden [2,3]. Diabetes diagnosis and care accounts for 6.3% of the UK’s health and social care budget, with 90% of all diabetes cases being T2D. Beyond the economic impact, T2D significantly affects people living with this condition. Most require lifelong disease management due to progressive hyperglycaemia, driven by worsening insulin resistance and declining beta-cell function [4]. Over time, chronic hyperglycaemia increases the risk of vascular complications and reduces life expectancy [5]. Early and effective glycaemic control is therefore a key goal of diabetes management and is associated with improved long-term outcomes [6,7].

Depression affects 18-25% of individuals with T2D [8] and is associated with worse diabetes outcomes, including a higher risk of complications [9] and earlier mortality [10]. It may therefore contribute to a more severe or accelerated T2D disease course [11]. Meta-analyses suggest that depression is associated with modest but clinically relevant increases in blood glucose levels [12,13], typically measured by glycated hemoglobin (HbA1c). However, this may underestimate its true effect on glycaemic control and disease progression in T2D as glucose-lowering treatment is typically intensified in response to rising HbA1c levels.

T2D is progressive [14], and this is evident in treatment patterns. In the UK, approximately 75% [15,16] of individuals with T2D are prescribed glucose-lowering medication (GLM). Treatment escalation from a single oral GLM (typically metformin) is common, occurring in 34-46% T2D patients in European healthcare settings [17–19]. Treatment progression, such as the addition or switching of GLMs, therefore offers a practical and clinically relevant way to assess the relationship between depression and the clinical course of T2D. Intensifying treatment is a clinical decision typically made in response to worsening glycemic control and may therefore reflect underlying disease progression. However, it can also be influenced by clinician and patient behaviours, or healthcare system factors. Rapid intensification may indicate accelerated disease progression and/ or timely care, while delays despite clinical need (clinical inertia) are associated with poorer outcomes.

Previous research has typically examined whether depression contributes to clinical inertia, but most findings do not support this. Studies generally report either no association [20–23] or higher rates [24–26] of treatment progression among individuals with depression. Results vary depending on the depression phenotype and study design used. Studies using depressive symptoms tend to report no association, whereas those using clinical diagnoses or antidepressant prescriptions suggest faster treatment progression. Few studies, however, have focused specifically on clinically coded depression within large, representative primary care populations. One prior study found that recent depression at T2D diagnosis, defined by recent hospital diagnosis or current antidepressant use, was associated with increased likelihood of GLM initiation [26]. This highlights the potential importance of depression recency in early T2D trajectories, but its relevance to ongoing treatment progression and diabetes management is unexplored. Moreover, no previous studies have explored recency while also considering individuals with more distant depression histories.

In this study, we used the large-scale Clinical Practice Research Datalink Aurum dataset [27] to assess the association between depression and T2D treatment progression, defined as treatment intensification and insulin initiation. Using flexible survival modelling, we investigated whether the timing of the most recent depression code prior to oral glucose-lowering monotherapy initiation influenced subsequent treatment progression. By incorporating depression recency, this study provides new insights into how comorbid mental health conditions may influence real-world diabetes management and clinical progression.

## 2. Research design and methods

### 2.1. Data sources

We conducted a retrospective cohort study using the December 2023 release of the Clinical Practice Research Datalink (CPRD) Aurum dataset [28], linked to Hospital Episode Statistics Admitted Patient Care (HES-APC) [29] and 2019 English Index of Multiple Deprivation (IMD) data. CPRD Aurum contains records for approximately 46.6 million individuals from 19.8% of primary care practices and includes demographics, diagnoses, prescriptions, and test results. Non-prescribing events, such as diagnoses, are recorded using CPRD medical codes mapped to SNOMED CT, Read, and local Egton Medical Information Systems (EMIS; software-specific) codes. Prescriptions are recorded using unique CPRD product codes mapped to Dictionary of Medicine and Devices (dm+d) codes. HES-APC provides inpatient care records from NHS hospitals in England, with diagnoses recorded using ICD-10 (international classification of diseases, 10^th^ revision) codes. Code lists used for cohort identification and variable definition are available online, with links provided in Supplementary Table 1.

### 2.2. Study population

The study included individuals with T2D, identified using Quality and Outcomes Framework (QOF) codes (Supplementary Materials), who met the following criteria: 1) T2D diagnosed after 01/04/2004 (T2D incorporated into the QOF), 2) aged ≥35 years at diagnosis, 3) registered at practice for ≥90 days prior to diagnosis, and 4) prescribed a single oral GLM (monotherapy) as their first ever GLM between 01/01/2011 and 31/12/2022.

Monotherapy medications considered were biguanides (metformin), sulfonylureas, dipeptidyl peptidase-4 inhibitors, sodium-glucose cotransporter 2 inhibitors, thiazolidinediones, alpha-glucosidase inhibitors (acarbose) and meglitinides. Oral semaglutide, a GLP-1 receptor agonist, was not considered as this is not currently a widely used first-line medication. T2D diagnosis date was defined as the earliest of: a T2D primary care code, HbA1c ≥48 mmol/mol (6.5%) or a GLM prescription.

We excluded individuals with: 1) fewer than two HbA1c measurements ≥48 mmol/mol (6.5%), or 2) non-T2D diabetes diagnoses including type 1, gestational, secondary or diabetes insipidus (Supplementary Materials). Merged practices were removed to prevent double counting.

The final study population included 378,935 individuals. The index date was monotherapy initiation, and patients could be followed for up to 12.75 years (01/01/2011 to 31/12/2023).

### 2.3. Outcomes

Two treatment progression outcomes were considered:

1. Time to treatment intensification: the first addition of a second GLM or a switch to a different GLM class. Changes in dose or formulation of the same medication were not considered. Individuals were followed from monotherapy initiation until the earliest of: treatment intensification, death, transfer out of the practice, last practice contribution date, study end date (31/12/2023), or six months after their last monotherapy prescription, to ensure follow-up only included periods when a change represented intensification rather than treatment re-start.
2. Time to insulin initiation: the first prescription of any insulin formulation. Follow-up extended from index until the earliest of: insulin initiation, death, transfer out of the practice, last date of practice data contribution, or the end of the study period (31/12/2023).

The time scale used in all models was years since monotherapy initiation.

### 2.4. Exposure (depression recency)

Depression was identified using primary care codes. Individuals were grouped according to the time between their most recent depression code and monotherapy initiation, using cut-offs based on the interquartile range (IQR) among those with prior depression. This resulted in four categories:

- Recent depression: ≤1.7 years before index (N = 23,765, 6.3%),
- Intermediate depression:1.7–12.8 years before index (N = 47,492, 12.5%),
- Distant depression: >12.8 years before index (N = 23,752, 6.3%),
- No prior depression: no depression code recorded before index (N = 282,926, 74.9%; reference group).

### 2.5. Covariates

We included demographic variables, clinical factors, healthcare utilisation and T2D-related complications recorded at or before the index date. Demographic variables were gender, age at index (years), year at index, ethnicity and practice-level Index of Multiple Deprivation (IMD) deciles. Ethnicity was categorised into five groups: “white”, “south Asian”, “black”, “mixed” and “other”. Clinical factors were: T2D disease duration (years), body mass index (BMI), HbA1c (glycated hemoglobin; mmol/mol), smoking status and alcohol use. T2D disease duration is the difference between T2D diagnosis and monotherapy initiation. BMI measurements were required to be recorded between two years prior to the index date and up to seven days afterwards. HbA1c measurements had to be recorded between six months prior to the index date and up to seven days afterwards. Smoking status was classified as ‘current smoker’, ‘ex-smoker’ and ‘non-smoker’ based on the most recent primary care smoking code recorded within the five years prior to the index date or up to seven days afterwards. Alcohol use was categorised as “none”, “within limits”, “excess” or “harmful” based on the highest level ever recorded prior to or within seven days of the index date. Healthcare utilisation was defined as the number of unique dates with coded clinical activity in CPRD Aurum during the 12 months prior to index. This measure reflects overall primary care activity, including consultations and test results, rather than general practitioner visits alone. To avoid overlap with the exposure, dates containing depression codes were excluded. Prescriptions were not included in this measure.

T2D-related complications were retinopathy, neuropathy, diabetic nephropathy, chronic kidney disease stage three or above (CKD3+), hypertension, ischaemic heart disease (IHD), heart failure, myocardial infarction, peripheral artery disease (PAD), stroke, transient ischaemic attack (TIA) and hospitalisation for heart failure (primary cause). Chronic liver disease (CLD) was also included as a relevant comorbidity due to its association with metabolic dysfunction and drug metabolism. In sensitivity analyses, we also considered a summary variable capturing the total number of T2D-related complications present at index.

Further details on cohort and variable creation, as well as data cleaning procedures, can be found in Supplementary Materials.

### 2.7. Statistical Analysis

Sample characteristics at monotherapy initiation were summarised by depression recency using medians and IQRs for continuous variables and counts and percentages for categorical variables. Results of chi-square tests (categorical variables) or Kruskal-Wallis tests (continuous variables) for differences across depression groups are provided in Supplementary Table 2. Kaplan-Meier (KM) curves were plotted for an initial description of time to treatment progression by depression group.

#### 2.7.1. Primary Analysis

We fitted Royston-Parmar flexible parametric survival models [30] to estimate the association between depression recency and time to treatment progression. Both proportional hazards (PH) and proportional odds (PO) specifications were evaluated. Model comparisons using the Bayesian Information Criterion (BIC) showed that the PO model provided the best fit for both outcomes (Supplementary Table 3). Results are therefore presented from the PO models as odds ratios (OR) with 95% confidence intervals (CIs). All models were adjusted for demographic variables, clinical factors, healthcare utilisation, and T2D-related complications. The PO assumption was assessed, and no evidence of non-proportionality was found (Supplementary Materials).

Missing data for ethnicity, BMI, HbA1c, smoking status, alcohol use, and IMD were handled using multiple imputation (MI) by chained equations [31], with 30 imputed datasets generated. Imputation models included all analysis variables, the event indicator, and the Nelson–Aalen cumulative baseline hazard [32], with additional clinical variables (such as systolic and diastolic blood pressure) included as auxiliary variables (listed in Supplementary Materials). Estimates were combined using Rubin’s rules, with p-values calculated using a *t*-distribution with adjusted degrees of freedom [33].

Standardised survival curves for each depression group were generated by averaging model-based survival predictions over a random subsample of 10,000 individuals. This approach provided population-level, covariate-adjusted comparisons of absolute risk by estimating the average predicted probability of remaining progression-free if all individuals in the sample had been assigned to the same depression group [34]. Risk differences and differences in restricted mean survival time, comparing each depression group with the no-depression reference, were then calculated at one, three, five, and 10 years. Estimates and standard errors were combined across imputations using Rubin’s rules.

#### 2.7.2. Sensitivity Analysis

Complete-case models were first fitted and compared with MI results. As findings were consistent, subsequent sensitivity analyses were conducted using complete-case data to reduce computational burden. Two sensitivity analyses were performed:

1. Complication count model. Individual diagnoses were replaced with a single count of T2D-related complications.
2. Three-month landmark analysis. To minimise misclassification from early treatment changes due to side effects or planned titration rather than treatment progression, individuals who experienced the outcome within the first three months after index were excluded. Time-to-event was measured from the three-month landmark onward.

#### 2.7.3. Software and code

All analyses were conducted in R version 4.3.2. KM curves were generated using the *survival* package [35]; Royston-Parmar models using *flexsurv* [36]; MI using *mice*, with pooling via *mitools* [37,38]; and plots using *ggplot2* [39]. R code for analysis is available at https://github.com/alexgillett/T2D_trtment_prog/.

## 3. Results

### 3.1. Descriptive statistics

Baseline characteristics of adults with T2D who initiated oral GLM monotherapy, stratified by depression recency, are presented in Table 1. Of the 378,935 individuals included, 74.9% (N = 283,926) had no prior depression code, and 93.6% initiated with metformin (Supplementary Table 2). Characteristics varied across depression categories. The recent depression group was youngest (median age 54.5 years), followed by intermediate (57.6 years), no depression (60.5 years), and distant depression (63.6 years). Women were more common in all depression groups (54.5–56.5%) compared with the no depression group (37.7%).

**Table 1.**
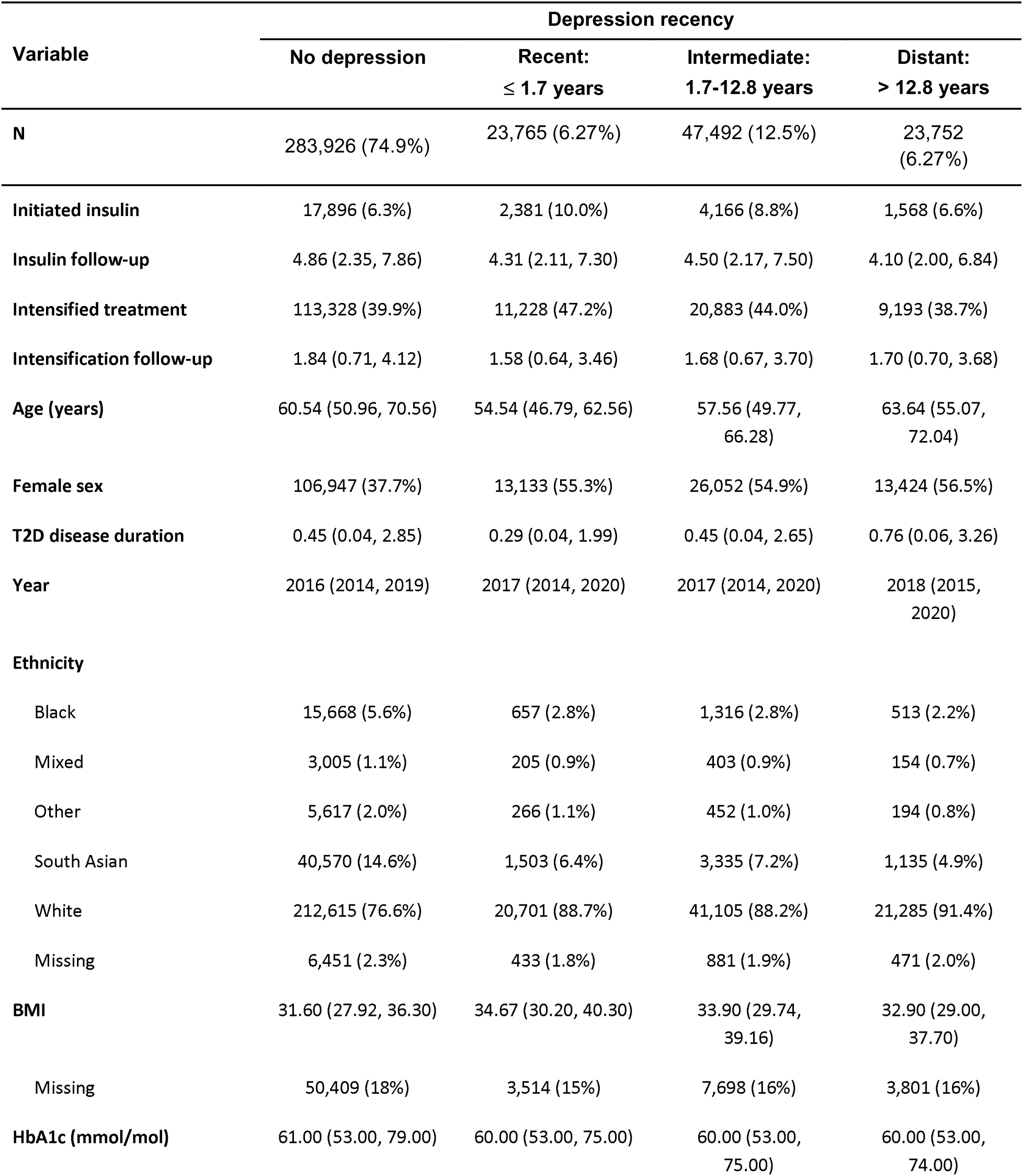
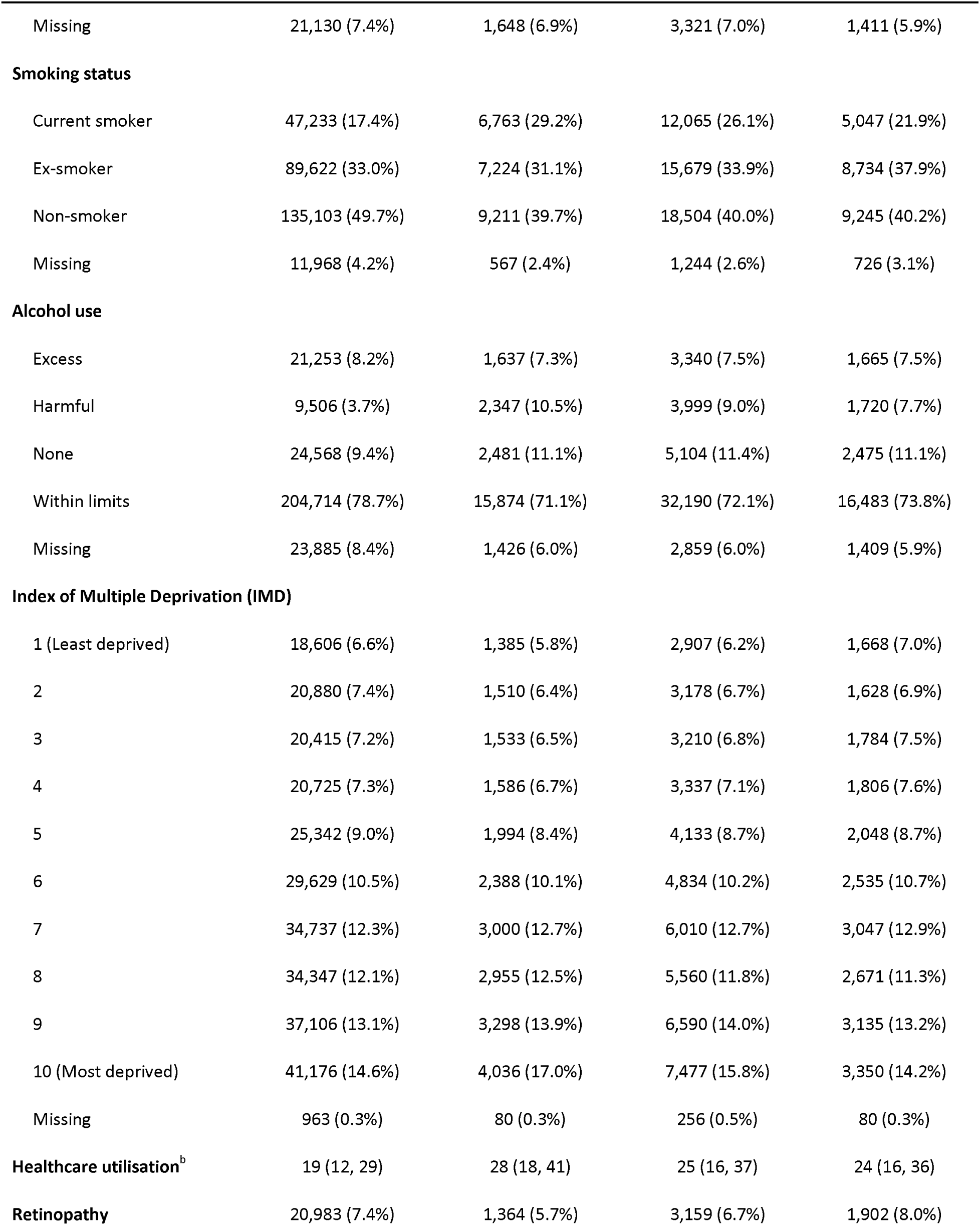
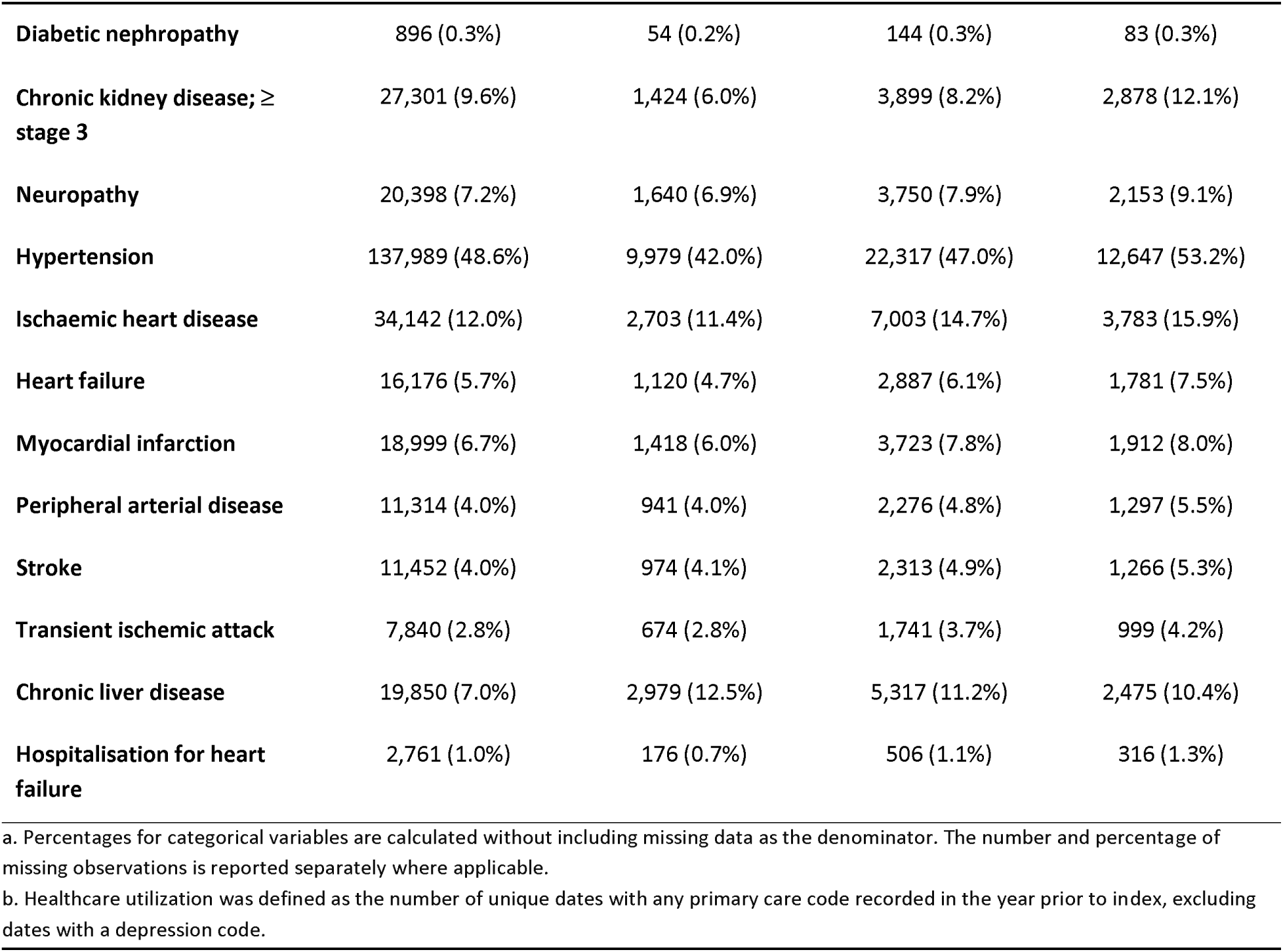
Participant characteristics at oral monotherapy initiation of glucose-lowering medication stratified by depression recency. Categorical variables^a^ are presented as counts and percentages, and continuous variables as medians and interquartile ranges (IQRs). Supplementary Table 2 provides p-values for between-group comparisons.

Health-related behaviours also differed. Current smoking was more prevalent among those with recent or intermediate depression (29.2% and 26.1%, respectively) than those without a depression history (17.4%), and harmful alcohol use followed a similar pattern. Healthcare utilisation was highest in the recent depression group (median 28 activity dates in the year prior to index, compared to 19 in the no-depression group). CLD was more common among individuals with a history of depression, particularly recent depression, while cardiovascular comorbidities (e.g., stroke, myocardial infarction, heart failure) were most common in the distant depression group, which also had the oldest participants on average.

Follow-up time was defined separately for each outcome as time from index to the event of interest or censoring. The median follow-up time for treatment intensification was 1.8 years (IQR: 0.7 - 4.0), during which 154,632 individuals (40.8%) added or switched GLM. For insulin initiation, median follow-up was 4.7 years (IQR: 2.3 - 7.7), with 26,011 individuals (6.9%) initiating insulin.

KM curves showed that, following monotherapy initiation, individuals with depression tend to experience both intensification and insulin initiation earlier than those without (Figure 1). The difference was most pronounced for recent depression and narrowed for distant depression.

**Figure 1.**
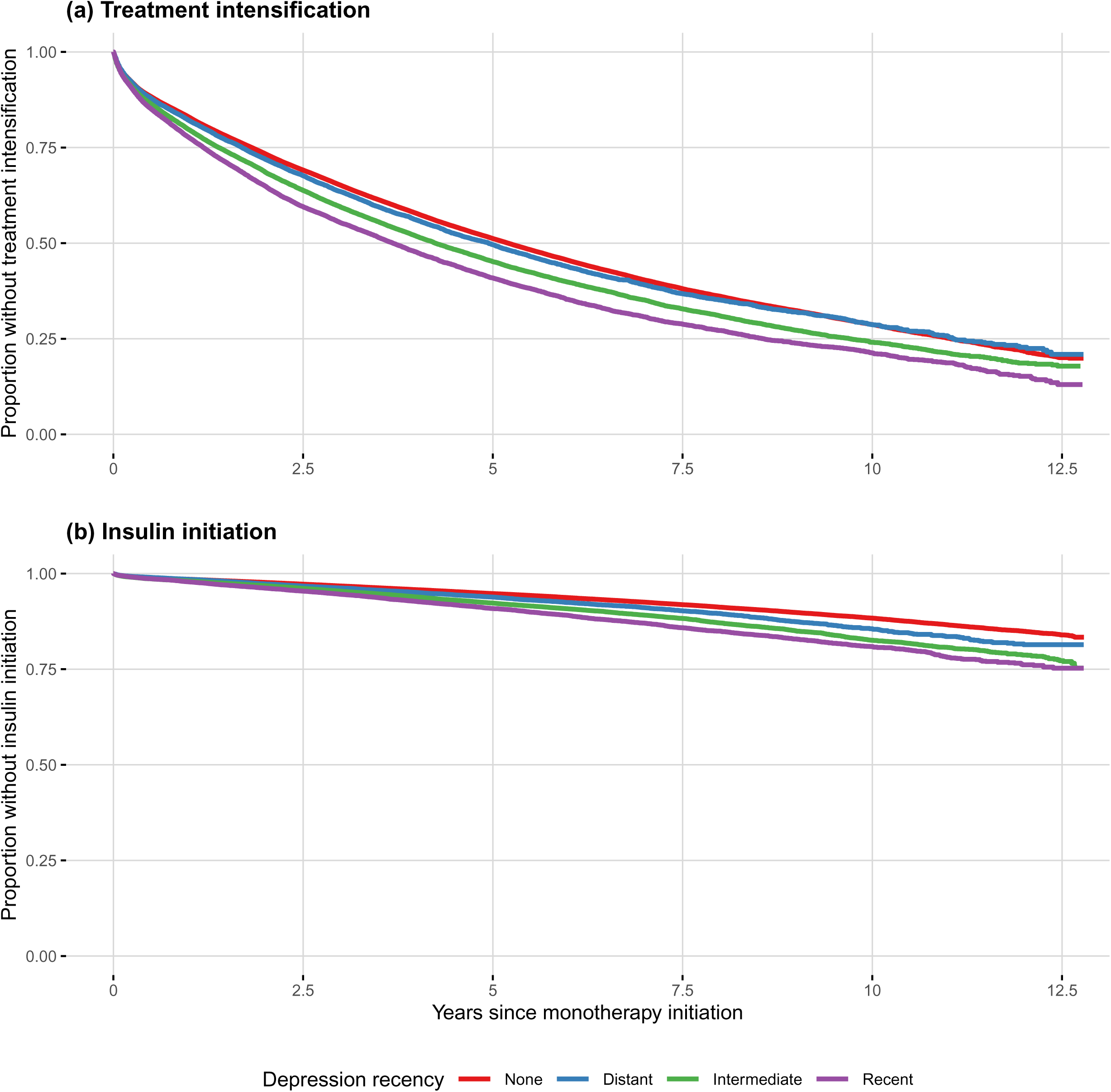
Kaplan-Meier curves for time to (a) treatment intensification and (b) insulin initiation, following initiation of oral glucose-lowering monotherapy, stratified by depression recency. Depression categories are defined as: none (no prior depression), distant (>12.8 years before index), intermediate (1.7-12.8 years) and recent (≤1.7 years).

### 3.2. Primary analysis of treatment intensification and insulin initiation

Individuals with a history of depression, particularly those with a more recent episode, had higher risks of treatment intensification and insulin initiation (Figure 2, Table 2, Supplementary Table 4). For treatment intensification, ORs were 1.20 (95% CI: 1.17-1.23) for recent depression, 1.12 (95% CI: 1.10-1.15) for intermediate, and 1.06 (95% CI: 1.03-1.09) for distant depression. For insulin initiation, ORs were 1.29 (95% CI: 1.23–1.36), 1.22 (95% CI: 1.18–1.27), and 1.11 (95% CI: 1.05–1.18) for recent, intermediate, and distant depression, respectively. Results were broadly consistent when using PH models (Supplementary Table 5), indicating robustness to survival model specification.

**Figure 2.**
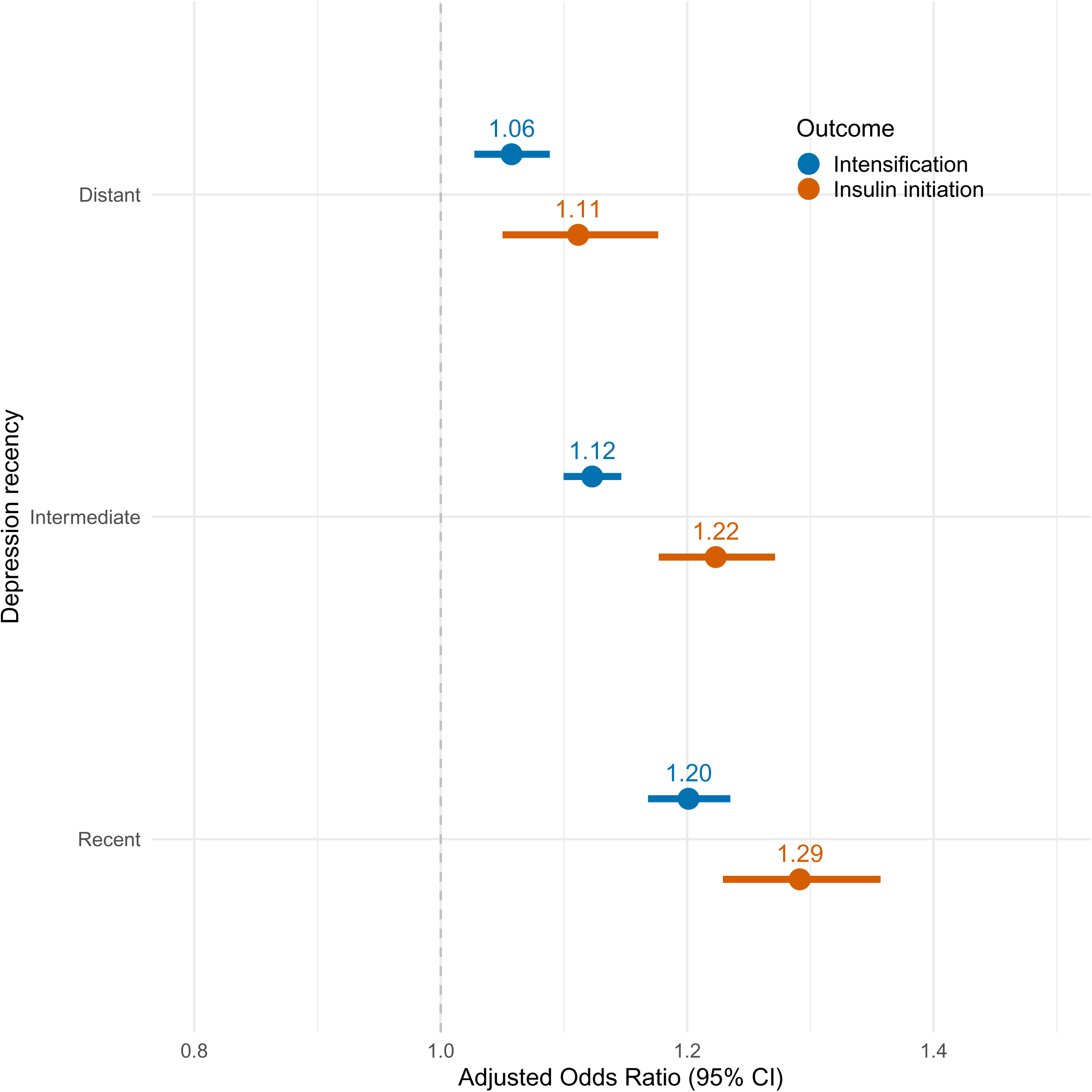
Association between depression recency and (1) treatment intensification (blue) and (2) insulin initiation (red), showing adjusted odds ratios with 95% confidence intervals (CIs). The reference group is no prior depression. Depression categories are defined as distant (>12.8 years before index), intermediate (1.7-12.8 years) and recent (≤1.7 years). Estimates are pooled across 30 imputed dataset.

**Table 2.** Association of depression recency with time to treatment intensification and time to insulin initiation. Results are from Royston-Parmar proportional odds models for a) the primary (multiple imputation) analysis, b) the complete case analysis (CC), and two sensitivity analyses based on the CC dataset: c) the complication count analysis (CC-count) and d) the three-month landmark analysis (CC-landmark).

| Analysis | N | Depression recency |  |  |  |  |  |  |  |  |
| --- | --- | --- | --- | --- | --- | --- | --- | --- | --- | --- |
|  |  | Recent: ≤ 1.7 years |  |  | Intermediate: 1.7-12.8 years |  |  | Distant: > 12.8 years |  |  |
|  |  | OR | 95% CI | p-value | OR | 95% CI | p-value | OR | 95% CI | p-value |
| Outcome: Time to treatment intensification |  |  |  |  |  |  |  |  |  |  |
| Primary | 378,935 | 1.20 | (1.17-1.23) | 3.05E-38 | 1.12 | (1.10-1.15) | 1.23E-27 | 1.06 | (1.03-1.09) | 1.53E-04 |
| CC | 268,197 | 1.21 | (1.17-1.25) | 4.58E-31 | 1.12 | (1.10-1.15) | 8.27E-21 | 1.06 | (1.02-1.09) | 1.23E-03 |
| CC-count | 268,197 | 1.21 | (1.17-1.25) | 4.42E-31 | 1.12 | (1.10-1.15) | 1.04E-20 | 1.06 | (1.02-1.09) | 1.37E-03 |
| CC-landmark | 246,701<sup>a</sup> | 1.20 | (1.16-1.24) | 4.41E-24 | 1.09 | (1.06-1.11) | 2.58E-09 | 1.06 | (1.02-1.10) | 3.64E-03 |
| Outcome: Time to insulin initiation |  |  |  |  |  |  |  |  |  |  |
| Primary | 378,935 | 1.29 | (1.23-1.36) | 4.05E-24 | 1.22 | (1.18-1.27) | 1.38E-24 | 1.11 | (1.05-1.18) | 2.59E-04 |
| CC | 268,197 | 1.30 | (1.23-1.38) | 3.79E-18 | 1.23 | (1.17-1.29) | 1.84E-17 | 1.11 | (1.03-1.19) | 4.26E-03 |
| CC-count | 268,197 | 1.30 | (1.22-1.38) | 7.53E-17 | 1.22 | (1.17-1.28) | 4.03E-17 | 1.11 | (1.03-1.19) | 4.05E-03 |
| CC-landmark | 264,282<sup>b</sup> | 1.35 | (1.27-1.43) | 3.14E-21 | 1.23 | (1.17-1.29) | 1.06E-16 | 1.12 | (1.05-1.21) | 1.52E-03 |
| a. Excluded participants who increased or added medications within the first three months post-index. b. Excluded participants who initiated insulin within the first three months post-index.<br>OR = odds ratio; CI = confidence interval. Exposure reference level = ‘None’ (no prior depression). |  |  |  |  |  |  |  |  |  |  |

Standardised survival analyses showed modest but higher absolute risks of progression for individuals with depression compared with no depression, with the largest differences in the recent depression group (Supplementary Table 6). Progression-free survival declined slightly faster in this group, indicating earlier treatment progression (Supplementary Figure 1). Pooled estimates of risk differences and differences in RMST at selected time points are shown in Table 3. Risk differences represent the percentage-point increase in progression for each depression subgroup compared with the no depression group, while differences in RMST quantifies how much earlier (in months) treatment progression occurred. At five years after monotherapy initiation, recent depression was associated with a 3.9% (95% CI: 3.4% - 4.5%) higher risk of treatment intensification and a 1.4% (95% CI: 1.1% – 1.7%) higher risk of insulin initiation compared to no depression, with intensification occurring on average two months earlier and insulin initiation about half a month earlier. These population-standardised estimates complement the relative effect measures (ORs) presented in Figure 2 by showing the side of absolute differences in treatment progression.

**Table 3.**
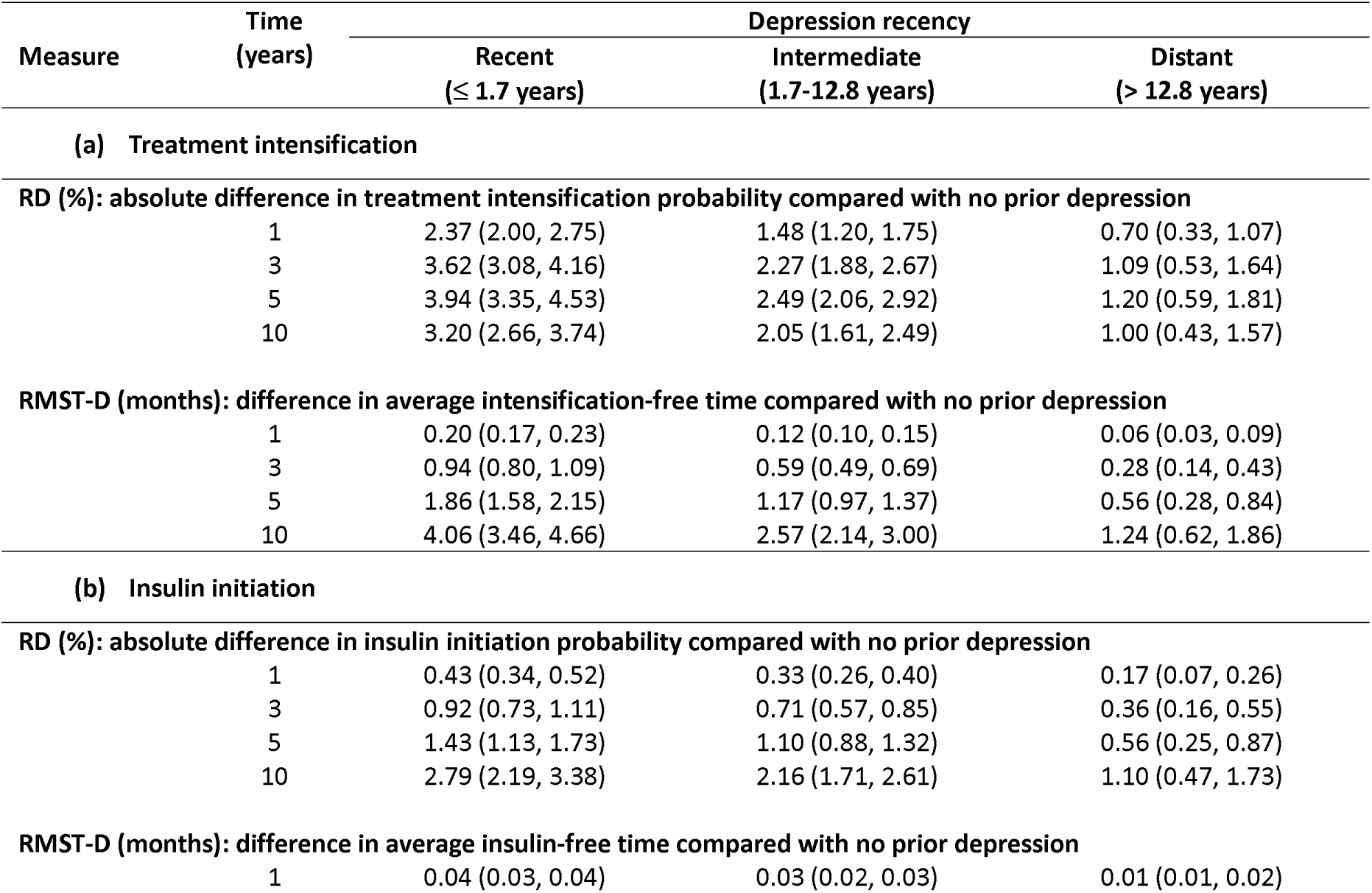
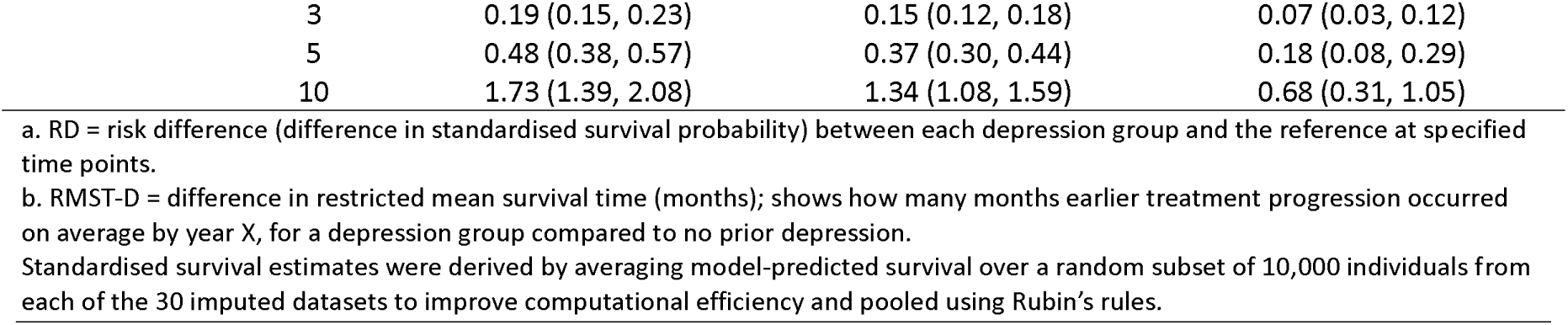
Absolute differences in treatment progression risk and progression-free time by depression recency. Risk differences (RD^a^; %) and differences in restricted mean survival times (RMST-D^b^; months), each with 95% confidence intervals, are presented for (a) treatment intensification and (b) insulin initiation, evaluated at 1-, 3-, 5-, and 10-years following monotherapy initiation (index). Positive values indicate higher progression risk (RD) or shorter progression-free time (RMST-D) compared with individuals with no prior depression (reference).

### 3.3. Sensitivity analyses

Complete-case analyses (N = 268,197; 70.8% of the MI sample) produced effect estimates of similar magnitude to those from the primary analysis, with all depression groups showing higher odds of T2D treatment progression compared to no depression (Table 2). As results were consistent, complete-case data were used for subsequent sensitivity analyses.

Modelling the number of diabetes-related complications as a count variable showed that a greater complication burden was associated with faster progression to both treatment intensification (OR 1.05, 95% CI: 1.04-1.06) and insulin initiation (OR 1.16, 95% CI: 1.14-1.17). However, model fit did not improve compared with the model including individual diagnoses, as indicated by the BIC (Supplementary Tables 7-8). Estimates for depression recency were largely unchanged (Table 2).

The landmark analysis, excluding individuals who intensified treatment within the first three months, produced results largely consistent with the MI analysis, suggesting that early progression did not unduly influence the observed associations (Table 2). ORs reduced slightly for the intermediate-recency depression group in the time to intensification model (1.12 to 1.09) and increased for recent depression in the insulin model (1.30 to 1.35).

Full model results for sensitivity analyses are provided in Supplementary Tables 7-8.

## 4. Discussion

In this large primary care cohort of 378,935 individuals with T2D initiating oral GLM monotherapy, a history of clinically diagnosed depression was associated with earlier treatment intensification (adding or switching to a new medication class) and insulin initiation. The effect was strongest for those with a more recent depression episode (within 20 months of monotherapy initiation), but was also observed in those with more distant depression histories. These findings were consistent across model specifications and sensitivity analyses.

Absolute differences in the risk of treatment progression were modest. For example, recent depression conferred a 3.9% higher risk of treatment intensification, and a 1.4% higher risk of insulin initiation over the first five years compared with those without depression. These differences correspond to approximately two months earlier treatment intensification and about half a month earlier insulin initiation. Although modest, these differences could translate to potentially large numbers of affected individuals at the population level, given the high prevalence of both depression and T2D. Our results highlight how relative and absolute measures provide complementary perspectives. Conditional ORs quantify relative associations after covariate adjustment, whereas standardised risk differences reflect population-average changes in progression probability. ORs were larger for insulin initiation compared to treatment intensification, but risk differences were smaller because insulin initiation occurred less frequently.

Depression was not associated with delayed treatment escalation, suggesting no evidence for clinical inertia. While we did not test inertia directly (e.g., time to treatment intensification after HbA1c exceeded 53 mmol/mol [7%]), our time-to-event models adjusted for baseline HbA1c and healthcare utilisation and consistently showed faster progression in those with depression. This pattern could reflect earlier glycaemic deterioration, more responsive diabetes care, or both. These findings are broadly consistent with previous research, although results have varied depending on the depression definition and study design [20–26]. Studies using current depressive symptoms (e.g., PHQ-9, HADS-D, or Edinburgh Depression Scale) typically report no association with time to insulin or treatment intensification [20–23]. In contrast, studies using depression diagnoses or antidepressant use show faster progression. A matched case-control study nested within CPRD and restricted to individuals with comorbid depression and T2D found that antidepressant use was associated with higher odds of insulin initiation, possibly reflecting greater depression severity among those prescribed antidepressants [25]. A large Danish cohort reported higher rates of GLM initiation among individuals with depression defined by recent hospital diagnosis or antidepressant use [26], and a US Veterans Health study found higher rates of treatment intensification following poor glycaemic control in those with depressive disorders [24].

This study extends previous work in several important ways. It is the largest to date (N = 378,935) to examine the association between clinically recorded depression and T2D treatment progression in a real-world UK primary care setting. Our rigorous statistical modelling applied flexible Royston-Parmar proportional odds models, which provided a better fit to the data than proportional hazard models. Analysis was anchored at initiation of oral GLM monotherapy, providing a consistent and clinically meaningful baseline for assessing treatment progression. This avoids the heterogeneity of designs that enrol participants at different stages of the diabetes care pathway. Finally, we incorporated depression recency to distinguish recent from more distant episodes.

Recent depression was strongly associated with treatment progression. This may reflect acute glycaemic effects, impaired self-management, or increased clinical vigilance. Depression can negatively impact medication adherence, diet, exercise, and sleep [40], and disruption in these behaviours can lead to worsening glycaemic control [41]. Additionally, depression is increasingly recognised as a risk factor for poor diabetes outcomes [42,43], which may prompt clinicians to monitor affected individuals more closely and act more swiftly when deterioration is detected. Supporting this, people with comorbid depression have been shown to undergo more frequent HbA1c testing [44], allowing earlier identification of poor glycaemic control.

Our results support the possibility that depression is linked to earlier and more rapid T2D progression. Although more responsive care may also impact treatment progression, our analysis adjusted for baseline HbA1c and primary care utilisation to account for initial glycaemic control and healthcare engagement. Depression confers higher risks of diabetes complications and early mortality [9,42,43], suggesting that timely care may not mitigate the adverse effects of comorbid depression. Contributing mechanisms may include behavioural factors and physiological pathways [40], such as chronic low-grade inflammation and dysregulation of the hypothalamic-pituitary-adrenal (HPA) axis leading to prolonged cortisol exposure [45,46].

Treatment progression is a complementary clinical marker of T2D disease course alongside HbA1c, which captures current glycaemic control but not the treatment intensity required to achieve it. Increases in GLMs required to maintain control are recognised indicators of progressive disease [47], yet the pace of intensification also depends on clinical decision-making, medication side effects, patient preferences, and clinical guidelines. Adverse outcomes are associated with both delayed intensification, which can lead to prolonged hyperglycaemia, and rapid intensification, which may indicate deteriorating glycaemic control despite therapy [48,49]. Treatment progression therefore remains a clinically meaningful outcome that reflects important changes in T2D [47].

Our findings show that clinical depression, particularly when recent, contributes to faster diabetes progression. Clinicians may be appropriately intensifying treatment in response to poorer glycaemic trajectories among people with depression, yet the persistence of elevated complication and mortality risks despite intervention raises concerns that conventional care may be insufficient. This underscores the potential value of integrated care models that address both mental and physical health in T2D management.

Several limitations should be noted. Residual confounding may remain from unmeasured factors such as depression severity, psychosocial stressors, or social support. Although models adjusted for baseline HbA1c, BMI, healthcare utilisation and T2D complications, reverse causation cannot be ruled out, particularly for recent depression, where worsening glycaemic control may itself contribute to depressive symptoms. The assumption of missing-at-random (MAR) in multiple imputation may not fully hold, though auxiliary information was included to mitigate this risk. Our definition of treatment progression excluded dose increases within the same medication class, potentially underestimating early treatment intensification. Additionally, not all depression episodes will be captured in our dataset, for example, due to not seeking care, receiving care outside of contributing practices or incomplete coding. Some individuals recorded as having no history or only distant depression may have experienced more recent or recurrent episodes. Depressive symptoms below diagnostic thresholds may also impact diabetes care and progression. We did not incorporate antidepressant prescribing, which can act as a proxy for more severe or persistent depression and may influence T2D progression directly through treatment-related weight gain and metabolic effects [50]. Finally, treatment progression reflects both the underlying disease trajectory and clinical decision-making, and may be influenced by prescriber preferences, treatment guidelines, patient preferences, and broader healthcare system factors.

### 4.1. Conclusion

Depression, particularly a more recent episode, is associated with earlier treatment intensification and insulin initiation in T2D. These findings highlight the importance of integrating mental health support into diabetes care.

## Supporting information

Supplementary Figure 1: Standardised survival curves

## Data Availability

All patient-level data in the present study are available to approved researchers upon request to CPRD. All summary-level data produced in the present work are contained in the manuscript and supplementary materials.

https://www.cprd.com/

https://github.com/alexgillett/T2D_trtment_prog/

## CRediT authorship contribution statement

**ACG:** Conceptualization, Funding acquisition, Data curation, Formal Analysis, Investigation, Methodology, Visualisation, Writing - original draft. **DH:** Data curation, Writing - review & editing. **RB:** Writing - review & editing. **KGY:** Data curation, Software, Writing - review & editing. **JT:** Conceptualization, Funding acquisition, Resources, Writing - review & editing. **CML:** Conceptualization, Funding acquisition, Supervision, Resources, Writing - review & editing.

## Availability of data and materials

This study used anonymised patient-level primary care data from the Clinical Practice Research Datalink (CPRD) Aurum, obtained under license from the UK Medicines and Healthcare products Regulatory Agency. The protocol was approved by the CPRD Independent Scientific Advisory Committee (protocol ID: 23_002544). Data access requests can be made to CPRD. Code for data management and analysis is available here: https://github.com/alexgillett/T2D_trtment_prog/.

## Declaration of competing interest

No potential or existing conflicts of interest relevant to this article were reported.

## Funding

This research is supported by the Medical Research Council (MR/X009815/1). This paper represents independent research part-funded by the National Institute for Health Research (NIHR) Maudsley Biomedical Research Centre at South London and Maudsley NHS Foundation Trust and King’s College London. The views expressed are those of the author(s) and not necessarily those of the NHS, the NIHR, or the Department of Health and Social Care.

## Acknowledgements

For the purpose of open access, the authors have applied a Creative Commons Attribution (CC BY) licence to any Author Accepted Manuscript version arising.

## Notes

### Competing Interest Statement

The authors have declared no competing interest.

### Author Declarations

This study used anonymised patient-level primary care data from the Clinical Practice Research Datalink (CPRD) Aurum (database version December 2023), obtained under license from the UK Medicines and Healthcare products Regulatory Agency. The protocol was approved by the CPRD Independent Scientific Advisory Committee (protocol ID: 23_002544).

