## Supplementary Materials for "Impact of depression on treatment progression in type 2 diabetes: A UK retrospective cohort study using the Clinical Practice Research Datalink Aurum database"

Gillett, *et al*

| Section | | Page |
| --- | --- | --- |
| 1. | Identifying individuals with type 2 diabetes and their diagnosis date | 1 |
| 2. | Defining the analysis cohort | 2 |
| 3. | Depression (exposure) extraction and cleaning | 3 |
| 4. | Covariates: details, extraction and cleaning | 4 |
| 5. | Brief description of Royston-Parmar proportional odds models | 8 |
| 6. | Violations of proportional odds assumption | 9 |

**1. Identifying individuals with type 2 diabetes and their diagnosis date**

Access to individuals with diabetes, identified using the ‘*all diabetes*’ code list (see Supplementary Table 1 for the code list link), was requested from CPRD Aurum. This list includes codes for type 1 diabetes (T1D), type 2 diabetes (T2D), gestational diabetes and gestational diabetes history, insulin receptor antibodies, malnutrition-related diabetes, maturity-onset diabetes of the young (MODY), secondary diabetes, other type, and non-specific diabetes codes.

Data was accessed through the multi-project licence of Exeter via protocol ID 23_002544 (<https://www.cprd.com/approved-studies/understanding-associations-between-major-depression-and-type-2-diabetes>). Data extraction, cleaning and cohort creation followed the established approach of the Exeter Diabetes Team. Links to relevant documentation are provided below; further details for the code lists and algorithms can be found at <https://github.com/Exeter-Diabetes/CPRD-Codelists/tree/pre-2024> and <https://github.com/Exeter-Diabetes> respectively.

From the *all diabetes* population, we retained individuals with at least one Quality and Outcomes Framework (QOF) diabetes code (Supplementary Table 1; *qof diabetes* code list link) recorded within valid follow-up. Valid follow-up is defined as being between each patient’s date of birth and data end date). We therefore derived:

- An approximate date of birth as follows: if both month and day were missing, we assigned 1st July of the recorded year; if the month was available but the exact day was not, we assigned the 15th of that month; finally, if the earliest clinical code in the observation record occurred earlier than this constructed date, we used that earlier date instead. And,
- A data end date as the earliest of: the last data collection date from the practice, date of patient deregistration or date of death as provided by CPRD.

We excluded individuals with codes indicating gestational diabetes/history, secondary diabetes, maturity-onset diabetes of the young (MODY), other/unspecified or genetic/syndromic types, insulin-receptor antibodies, malnutrition-related diabetes, and any diabetes insipidus code (*diabetes exclusion* code list link in Supplementary Table 1).

For each patient we derived a diagnosis date as the earliest of: a diabetes code, HbA1c ≥48 mmol/mol (with unit cleaning and % to mmol/mol conversion when values ≤20; see Section 4 below for details), or a glucose-lowering medication (GLM) prescription.

Diabetes type (T1D versus T2D) was assigned using a rule-based algorithm (<https://github.com/Exeter-Diabetes/CPRD-Cohort-scripts/blob/main/all_diabetes_cohort.R>; <https://github.com/Exeter-Diabetes/CPRD-Codelists/blob/pre-2024/readme.md#defining-diabetes-type>) incorporating:

a) Insulin exposure: Patients with no record of any insulin prescription were classified as T2D by default. Patients who ever received insulin entered a further classification step based on their diagnosis codes and clinical profile.
b) Counts of diabetes type-specific codes: For insulin-treated patients, diabetes type codes in their record were counted. If multiple type-specific codes were present (e.g. some codes indicating T1D and some T2D), the predominant code type determined classification. Specifically, if T1D codes were at least double the number of T2D codes, the patient was classified as T1D; otherwise, they were classified as T2D. If exactly one type-specific diagnosis code was recorded, the patient was simply classified according to the single code type.
c) Age at diagnosis and time to insulin: If an insulin-treated patient had no T1D or T2D codes (only non-specific diabetes codes), the algorithm used age and insulin timing criteria. Patients diagnosed before age 35 who started insulin within 1 year of diagnosis were classified as T1D, whereas those who did not meet this early-insulin criteria were classified as T2D. Note, as described in Section 2 below, we excluded individuals diagnosed with T2D before the age of 35 (to reduce the risk of T2D versus T1D mis-classification). This exclusion also ensures that our analysis of the time to insulin initiation is biased by insulin-use within the first year being part of diabetes-type classification algorithm.
d) Current use of non-insulin GLM: In cases where the interval from diagnosis to insulin initiation could not be calculated (e.g. if diagnosis pre-dated the patient’s CPRD registration), patients diagnosed before age 35 with no current non-insulin GLM use were classified as T1D; all others were classified as T2D.

To minimise artefacts, particularly from diabetes codes recorded in a patient’s year of birth (YOB), classification was repeated after excluding these codes. Patients whose classification differed between the two runs were excluded as unclassifiable, and patients classified as T2D whose diagnosis date still fell in their year of birth after this step were also excluded.

Those classified as T1D were then removed. Individuals with T2D diagnosis dates prior to, or within 90 days after, GP registration start date were further excluded.

**2. Defining the analysis cohort**

Using the individuals identified with T2D via the approach outline in Section 1, we further excluded individuals if:

1) their T2D diagnosis date was before 1^st^ April, 2004 (when T2D was incorporated into the QOF),

2) they are aged < 35 years at diagnosis,

3) they were not prescribed a single oral GLM (monotherapy) as their first ever GLM between 01/04/2011 and 31/12/2022,

4) they had less than two HbA1c measurements ≥48 mmol/mol (6.5%) on record (indicating limited evidence of T2D), and

5) were from a merged practice (to prevent double counting).

The index date was then the date of monotherapy initiation.

**3. Depression (exposure) extraction and cleaning**

The code list used to identify individuals with depression is provided in Supplementary Table 1. Individual with any depression code recorded prior to the index date were classified as having a history of depression. For these individuals, the last recorded depression code prior to index date was identified, and the time interval between this last depression code and the index date was calculated. The distribution of this interval is shown in Supplementary Material Figure A. Quartiles of the distribution were then used to derive a categorical variable for depression recency.

**Supplementary Material Figure A.** Distribution of the time since last recorded depression code (years). Dashed vertical lines represent the interquartile range (25^th^ percentile = 1.7 years; 75^th^ percentile = 12.8 years) which were the cut points used to define depression recency.

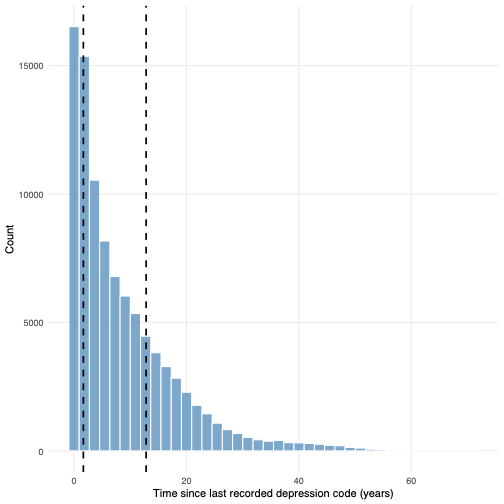

**4. Covariates: details, extraction and cleaning**

Code lists for covariates are all available online. Links to code lists are provided in Supplementary Table 1. For codes from primary care, provided code lists are either CPRD Aurum medical codes (medcodes) or product codes (prodcodes). Medcodes map to SNOMED CT, Read, and local Egton Medical Information Systems (EMIS; software-specific) codes via the CPRD medical code dictionary. Prodcodes map to Dictionary of Medicine and Devices (dm+d) codes via the product code dictionary. Please contact CPRD for information about accessing data dictionaries. HES-APC provides inpatient care records from NHS hospitals in England, with diagnoses recorded using ICD-10 (international classification of diseases, 10^th^ revision) codes, and operations, procedures and interventions recorded using OPCS-4 codes.

***List of main analysis covariates***

Gender, age at index, calendar year at index, T2D disease duration at index, ethnicity, practice level Index of Multiple Deprivation, smoking status at index, alcohol use at index, body mass index at index, HbA1c at index, healthcare utilization in the year before index, retinopathy, neuropathy, diabetic neuropathy, hypertension, ischaemic heart disease, heart failure, myocardial infarction, peripheral artery disease, stroke, transient ischaemic attack, hospitalisation for heart failure (primary cause), chronic kidney disease (CKD) stage 3 or above (CKD3+), and chronic liver disease.

***List of auxiliary covariates***

Systolic blood pressure, diastolic blood pressure, chronic obstruction pulmonary disease, bronchiectasis, asthma, pulmonary fibrosis, pulmonary hypertension, solid cancer, haematological cancer, solid organ transplant, rheumatoid arthritis, dementia, and family history of premature cardiovascular disease.

***List of sensitivity analysis covariates***

Count of main analysis T2D complications.

***Extraction and cleaning process***

**Gender:** Provided by CPRD in the primary care patient table. Gender is recorded as ‘male’, ‘female’ or ‘other’. Only the most recently recorded gender is provided. The category ‘other’ is rarely used by GPs in practice. No individuals in the analysis sample had their gender recorded as ‘other’.

**Age at index:** Difference in years between date of birth and date of monotherapy initiation. See Section 1 for how date of birth was defined.

**Year at index:** The year at which monotherapy was initiated. The median year at index was 2017. We centred the variable around this median.

**T2D disease duration at index:** Time between date of T2D diagnosis and date of monotherapy initiation (years).

**Ethnicity:** An algorithm developed by Exeter university (https://github.com/Exeter-Diabetes/CPRD-Codelists/blob/main/readme.md#ethnicity) was used to generate a five-level ethnicity covariate. Briefly, the algorithm selects the most frequently recorded ethnicity code (excluding ‘unknown’). If multiple codes were equally frequent, the most recent was used. If multiple ethnicities were recorded on the most recent date, ethnicity was set to unknown. Where primary care ethnicity was missing, ethnicity from linked Hospital Episode Statistics (HES; provided in the HES patient table) was used if consistent with the 5-category definition.

The 5-category classification groups individuals as:

0 = White (British, Irish, Other White),

1 = South Asian (Indian, Pakistani, Bangladeshi, and Other Asian),

2 = Black (Caribbean, African, Other Black),

3 = Other (Chinese, Other Ethnic group),

4 = Mixed (White and Black Caribbean, White and Black African, White and Asian, Other Mixed),

5 = Unknown (Not Stated/ Unknown).

Brackets contain the GP categories extracted from CPRD primary care records.

**Practice-level index of multiple deprivation (IMD):** Postcode from practices were linked to the 2019 English Index of Multiple Deprivation (IMD), assigned at the Lower Layer Super Output Area (LSOA) surrounding the practice postcode (mean population ~1,600). The IMD is a composite relative deprivation measure combining income, employment, education, health, housing, crime, access to services, and living environment. Scores are reported in deciles and utilized as a ranked indicator of area-level deprivation, with 1 being the least deprived and 10 the most. This score is provided by CPRD (linkage to small area level data).

**Smoking status at index:** This variable was defined using the most recent smoking code within 5 years prior to index. Each individual was initially assigned to a QRISK2-aligned smoking category (0=non-smoker, 1=ex-smoker, 2=light smoker [1-10 cig/day], 3=moderate smoker [11-19 cig/day], 4=heavy smoker [20+ cig/day]). If both non-smoker and ex-smoker codes were recorded, then ex-smoker was selected. Where smoking codes included an associated numeric value (e.g. number of cigarettes per day), this value was used to refine categorisation, except for codes related to pack-years (e.g. medcode 1780396011), which were ignored. In cases where conflicting categories (e.g. non-smoker and current smoker) were recorded on the same date, the lowest (i.e. less intensive) category was selected. See: <https://github.com/Exeter-Diabetes/CPRD-Cohort-scripts/blob/main/template_smoking.R> and <https://github.com/Exeter-Diabetes/CPRD-Codelists#smoking> for R-code and further information.

After being assigned to a QRISK2-aligned smoking category we merged light-, moderate- and heavy- smokers into a single ‘current smoker’ category. This gave three categories overall: non-smoker, ex-smoker and current smoker.

**Alcohol use at index:** Individuals were assigned the most recently recorded alcohol consumption level, with the highest level selected if multiple values were recorded on the same date. If a level 3 (‘harmful’) code was ever recorded, the individual was classified as harmful drinker, regardless of subsequent codes. Categories were:

0 = None,

1 = Within limits,

2 = Excess,

3 = Harmful.

See <https://github.com/Exeter-Diabetes/CPRD-Cohort-scripts/blob/main/template_alcohol.R> and <https://github.com/Exeter-Diabetes/CPRD-Codelists?tab=readme-ov-file#alcohol-consumption> for R-code and further information.

**BMI at index:** If coded BMI values were available then these were preferentially used. If unavailable, BMI was calculated using weight and height measurements recorded on the same day. Implausible values were excluded using the following plausible ranges:

- BMI: 15-100 kg/m^2^
- Weight: 40-350 kg
- Height: 60-225 cm

We then restricted to BMI values recorded within the two years before and up to 7 days after index date, selecting the value to closest to index.

**HbA1c at index (mmol/mol):** Measurements recorded before 1990 were excluded, as HbA1c was not routinely used in UK primary care before this time. HbA1c values ≤20 were assumed to be recorded in percentage units and were converted to mmol/mol using the standard NGSP-to-IFCC formula. Values outside the plausible range (20–195 mmol/mol) were then removed. To capture glycaemic control at oral monotherapy initiation (index), we restricted to HbA1c measurements recorded within six months prior to and up to seven days after the index date, selecting the value closest to index.

**Healthcare utilisation:** Healthcare utilisation was defined as the number of unique dates with medical code activity in CPRD Aurum during the 12 months prior to index (i.e., in the CPRD observation table). This measure reflects overall primary care activity, including consultations and test results, rather than general practitioner (GP) visits alone. To avoid overlap with the exposure, dates containing depression codes were excluded. Prescriptions were not included in this measure.

**T2D-related comorbidities and complications (measured at index)**

A binary variable indicating whether a complication or comorbidity was present at index or not was created for the following:

- **Primary analysis covariates:** retinopathy, neuropathy, diabetic neuropathy, hypertension, ischaemic heart disease, heart failure, myocardial infarction, peripheral artery disease, stroke, transient ischaemic attack, hospitalisation for heart failure (primary cause), chronic kidney disease (CKD) stage 3 or above (CKD3+), and chronic liver disease.
- **Auxiliary variables used in multiple imputation:** chronic obstruction pulmonary disease, bronchiectasis, asthma, pulmonary fibrosis, pulmonary hypertension, solid cancer, haematological cancer, solid organ transplant, rheumatoid arthritis, dementia, and family history of premature cardiovascular disease.

For each complication/ comorbidity, we compiled diagnoses and procedures recorded in CPRD (medcodes), HES-APC (ICD-10 diagnoses), and HES-APC/OPCS-4 (procedures). All codes were cleaned to remove records before a patient’s date of birth or after censoring (death, transfer out, or last collection date). We then linked these occurrences to each patient’s index date (first oral GLM monotherapy prescription).

A complication/ comorbidity was considered present at index if the patient had at least one relevant code recorded on or before the index date. Relevant code lists are provided in Supplementary Table 1.

**Note 1:** Hospitalisation for heart failure (a primary analysis covariate) was defined only from HES-APC ICD-10 codes for heart failure, restricting to episodes where heart failure was the primary (first-position) diagnosis. All other steps were as described above.

**Note 2:** Hypertension was only defined using primary care records, not link HES records.

**Note 3:** **Chronic kidney disease stage 3 and above (CKD3+) at index** extraction requires further description. The date at which each CKD stage was first observed was derived using an algorithm developed by the Exeter Diabetes group. Briefly, estimated glomerular filtration rate (eGFR) was calculated from serum creatinine using the CKD-EPI 2021 equation (<https://www.kidney.org/ckd-epi-creatinine-equation-2021-0>). CKD stage was assigned according to eGFR thresholds (stage 1: ≥90; stage 2: 60–89; stage 3: 30–59; stage 4: 15–29; stage 5: <15 ml/min/1.73 m²). To distinguish chronic disease from transient changes, CKD stages 3–5 were only assigned if ≥2 qualifying eGFR values were observed at least 90 days apart. Patients with diagnostic or procedure codes indicating end-stage kidney disease (dialysis, transplant, stage 5 CKD) prior to index were also classified as CKD3+. For the purposes of the analysis, CKD3+ was coded as a binary variable (present/absent at index).

For more details on the algorithm please see <https://github.com/Exeter-Diabetes/CPRD-Codelists/tree/pre-2024?tab=readme-ov-file#ckd-chronic-kidney-disease-stage> for a description, and <https://github.com/Exeter-Diabetes/CPRD-Cohort-scripts/blob/Oct2020-download/all_patid_ckd_stages.R> and <https://github.com/Exeter-Diabetes/CPRD-Cohort-scripts/blob/Oct2020-download/template_ckd_stages.R> for the R- code. Supplementary Table 1 contains code lists for the required variables (creatinine, CKD diagnostic codes).

**Additional variables- sensitivity analyses**

**Number of T2D-related complications present at index:** A count of the number of complications/ comorbidities used in the primary analysis (see above) that an individual had recorded at index.

**Additional variables- further auxiliary variables used in multiple imputation (measured at index)**

**Systolic blood pressure (SBP; mmHg):** SBP measurements were extracted from CPRD Aurum using codes provided in Supplementary Table 1. Implausible values were excluded using the following:

- <40 mmHg and
- >270 mmHg.

If multiple measurements were recorded on the same day, their mean was taken. We restricted to measurements taken within two years before up to 7 days after the index date and selected the measurement closest to index.

**Diastolic blood pressure (DBP; mmHg):** DBP measurements were extracted from CPRD Aurum using codes provided in Supplementary Table 1. Implausible values were excluded using the following:

- <30 mmHg and
- >200 mmHg.

If multiple measurements were recorded on the same day, their mean was taken. We restricted to measurements taken within two years before up to 7 days after the index date and selected the measurement closest to index.

**Stage 5 Chronic Kidney Disease (CKD5):** End stage renal disease defined using diagnostic codes from primary or secondary care for stage 5 CKD, recorded at or before the index date.

**5. Brief description of Royston-Parmar proportional odds models**

Primary time-to-event analyses were conducted using Royston-Parmar proportional odds (RP-PO) models. These models estimate the log cumulative odds of failure as a smooth function of log-time using restricted cubic splines, plus a linear predictor of covariates. The PO specification assumes that covariate effects are constant over time on the log cumulative odds scale (akin to the proportional hazards assumption in Cox models).

For the main analyses, models with five internal spline knots (k=5) were fitted.

Let $T$ denote the time to event with cumulative distribution: $F(t|\underline{x}) = p(T\leq t|\underline{x})$

And survival function: $S(t|\underline{x}) = 1 - F(t|\underline{x})$

Then the RP-PO model is:

$$log\left( \frac{F\left( t | \underline{x} \right)}{S\left( t | \underline{x} \right)} \right) = log\left( \frac{1-S\left( t | \underline{x} \right)}{S\left( t | \underline{x} \right)} \right)$$

$$= h\left( log\left( t \right);\underline{\gamma} \right) + \underline{x}^{T}\underline{\beta}$$

Where $h\left( log\left( t \right); \underline{\gamma} \right)$ is a restricted cubic spline (RCS) function of $log\left( t \right)$ with coefficients $\underline{\gamma}=\left( \gamma_{0},\gamma_{1},...,\gamma_{k+1} \right)$and:

$$h\left( log\left( t \right);\underline{\gamma} \right)=\gamma_{0} +\gamma_{1}log\left( t \right)+\gamma_{2}\nu_{1}\left( log\left( t \right) \right)+...+\gamma_{k+1}\nu_{k}\left( log\left( t \right) \right)$$

where the *j^th^* basis function is:

$$\nu_{k}\left( \theta\right) = \left( \theta-m_{j} \right)_{+}^{3}+\lambda_{j}\left( \theta-m_{min} \right)_{+}^{3}+\left( 1 - \lambda_{j} \right)\left( \theta-m_{max} \right)_{+}^{3}$$

for $j=1, 2, ..., k$, where:

- $\theta=log\left( t \right)$.
- $m_{min}$ and $m_{max}$ are boundary knots, typically placed at the most extreme observed $\theta$-values. Spline functions are constrained to be linear beyond these values.
- There are *k* internal knots in addition to $m_{min}$ and $m_{max}$ such that $m_{min}<m_{1}<m_{2}<...<m_{k}<m_{max}$.
- $\lambda_{j}=\left( m_{max}-m_{j} \right)/\left( m_{max}-m_{min} \right)$.
- $\left( \theta- a \right)_{+}=max(0, \theta- a)$.

Then the odds ratio (OR) associated with a +1 unit change in covariate $x_{i}$ is:

$${OR}_{i}= e^{\beta_{i}}$$

And is constant over time under the PO assumption.

**6. Violations of proportional odds assumption**

***Non-proportional (time-varying) effects***

To assess whether the association between depression recency and treatment progression changed over time, spline-based interactions with log-time were added to the PO models. Two models were considered:

- M1. Included an interaction between exposure and first spline function of log-time.
- M2. Extended M1 by further adding an interaction between exposure and the second spline function of log-time.

Model fit was compared using BIC. Standardised survival probabilities and risk differences were contrasted to assess the impact of allowing depression effects to vary over time.

***Model specification for the time varying extension***

To relax the PO assumption for a covariate $z$, time-varying effects can be introduced by interacting $z$ with the RCS basis functions. Then the model becomes:

$$log\left( \frac{1-S\left( t | \underline{x} \right)}{S\left( t | \underline{x} \right)} \right)= h\left( log\left( t \right);\underline{\gamma} \right) + \underline{x}^{T}\underline{\beta}+h\left( log\left( t \right);\underline{\delta} \right)z$$

To investigate potential non-proportional odds for depression over time we fitted two models using the complete case dataset. Model 1 (M1) contained an interaction between the first log-time spline basis (which is simply log-time) and depression recency. Model 2 extends M1 to further include an interaction between the second spline basis function and depression recency. That is:

**M1.** $log\left( \frac{1-S\left( t | \underline{x} \right)}{S\left( t | \underline{x} \right)} \right)= h\left( log\left( t \right);\underline{\gamma} \right) + \underline{x}^{T}\underline{\beta}+ \underline{z}^{T}\underline{\delta}_{1}log(t)$

**M2**. $log\left( \frac{1-S\left( t | \underline{x} \right)}{S\left( t | \underline{x} \right)} \right)= h\left( log\left( t \right);\underline{\gamma} \right) + \underline{x}^{T}\underline{\beta}+ \underline{z}^{T}\underline{\delta}_{1}log(t)+\underline{z}^{T}\underline{\delta}_{2}\nu_{1}\left( log\left( t \right) \right)$

Where:

- $\underline{z} =(z_{1}, z_{2},z_{3})$ contains the three dummy variables for depression recency.
- $\underline{\delta}_{1}= (\delta_{11}, \delta_{12},\delta_{13})$ are the parameters to capture the time-varying depression recency effects with respect to log(t).
- $\underline{\delta}_{2}= (\delta_{21}, \delta_{22},\delta_{23})$ are the parameters to capture the time-varying depression recency effects with respect to $\nu_{1}\left( log\left( t \right) \right)$.

***Results: No evidence of PO violations for depression exposure***

Compared with the PO model, the time-varying models (M1 and M2) provided minimal or no improvement in fit based on the BIC (Supplementary Tables 9a-10a). Consistent standardised survival estimates across models (Supplementary Tables 9b-10b) further indicated little evidence for violations of the PO assumption.
