## Supplementary Figure 1: Standardised survival curves for "Impact of depression on treatment progression in type 2 diabetes: A UK retrospective cohort study using the Clinical Practice Research Datalink Aurum database"

**Supplementary Figure 1.** Pooled, risk-standardised survival curves by depression recency for (a) treatment intensification, and (b) insulin initiation, estimated from Royston-Parmar proportional odds models pooled across 30 imputed datasets. Shaded areas show 95% confidence intervals.

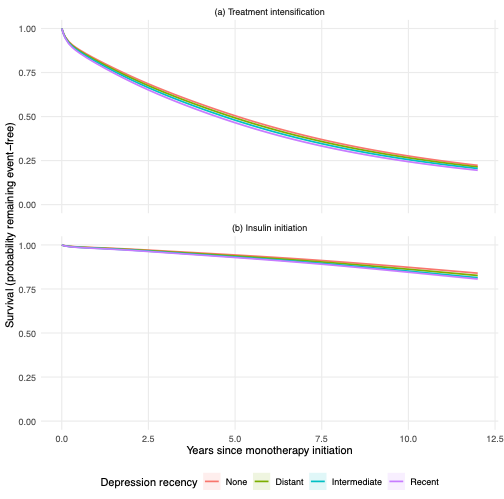
